# Two-Year Evaluation of *Legionella* in an Aging Residential Building: Assessment of Multiple Potable Water Remediation Approaches

**DOI:** 10.1101/2023.07.19.23292444

**Authors:** Monica Lee-Masi, Caroline Coulter, Steven J. Chow, Benjamin Zaitchik, Joseph G. Jacangelo, Natalie G. Exum, Kellogg J. Schwab

## Abstract

*Legionella* is an opportunistic waterborne pathogen that is difficult to eradicate in colonized drinking water pipes. *Legionella* control is further challenged by aging water infrastructure and lack of evidence-based guidance for building treatment. This study assessed multiple premise water remediation approaches designed to reduce *Legionella pneumophila (Lp)* within a residential building located in an aging, urban drinking water system over a two-year period. Samples (n=745) were collected from hot and cold-water lines and quantified via most probable number culture. Building-level treatment approaches included three single heat shocks (HS), three single chemical shocks (CS), and continuous low-level chemical disinfection (CCD) in the potable water system. The building was highly colonized with *Lp* with 71% *Lp* positivity. Single HS had a statistically significant *Lp* reduction one day post treatment but no significant *Lp* reduction one, two, and four weeks post treatment. The first two CS resulted in statistically significant *Lp* reduction at two days and four weeks post treatment, but there was a significant *Lp* increase at four weeks following the third CS. CCD resulted in statistically significant *Lp* reduction ten weeks post treatment implementation. This demonstrates that in a building highly colonized with *Lp*, sustained remediation is best achieved using CCD.

**SYNOPSIS:** Long-term *Legionell*a control is difficult to maintain within aging premise plumbing. This study supports continuous low-level building treatment as an effective long-term remediation of a building highly colonized with *Legionella*.

**For Table of Contents Only:** 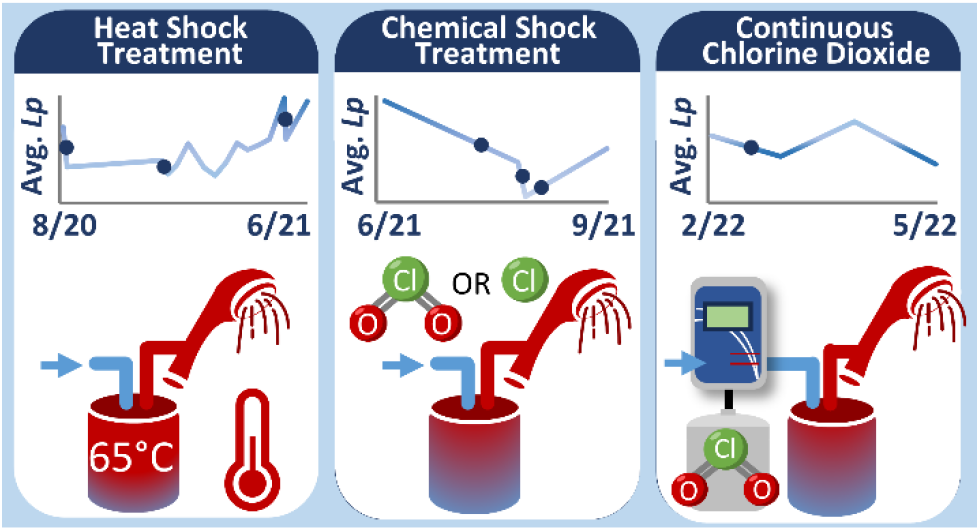

## INTRODUCTION

*Legionella* bacteria, the cause of Legionnaires’ Disease, is the leading cause of waterborne disease outbreaks attributed to drinking water in the United States (US).^1^ Legionnaires’ Disease is a severe form of pneumonia with a 10% case fatality rate in the general population and 25% case fatality rate in health-care settings.^2^ The Centers of Disease Control and Prevention (CDC) reported 10,000 cases of Legionnaires’ Disease in 2018, however the actual number of cases is estimated to be ∼1.8-2.7-fold higher.^1, 3^ While overall waterborne disease outbreaks in community drinking water systems have decreased over past decades due to improved drinking water regulations developed in the US Environmental Protection Agency (EPA) Safe Drinking Water Act, the number of reported Legionnaires’ Disease cases has risen 5.5-fold since the year 2000.^1, 3^ The cause of the increasing incidence is multi-factorial, with the aging drinking water infrastructure (building water systems and engineered water systems) playing a major role.^4, 5^

The majority of Legionnaires’ Disease outbreaks have been reported from buildings with complex plumbing systems, with infection occurring via inhalation of contaminated water from engineered water systems (e.g., cooling towers, air conditioners, taps and showers).^6^ *Legionella pneumophila* commonly colonize in hot water systems with human aerosolization exposure through faucets and showerheads.^7, 8^ *L. pneumophila* thrive in areas with warm temperatures (25-45 °C), low-water flow, and within biofilms, conditions which are characteristic to premise plumbing systems.^5^ Over time, aging pipes and plumbing systems can experience deficiencies, including leaks or corrosion, that lead to temperature fluctuations in pipes, water stagnation, and increased biofilm formation.^9^ These conditions can create an even more favorable environment for *L. pneumophila* growth and proliferation.^9^

Controlling for *Legionella* in premise plumbing systems can be a difficult and expensive undertaking.^5^ Currently there is no US federal regulation for *Legionella* monitoring in drinking water within premise plumbing.^10^ The CDC and EPA have developed guidelines for building managers and owners for preventing and controlling *Legionella* growth.^11, 12^ These guidelines include a comprehensive framework for developing and implementing water management programs (WMP) in buildings and treatment approaches.^11, 13^ A review for controlling *Legionella* bacteria in building systems found that the implementation of WMPs and periodic monitoring was the most effective approach to prevent Legionnaires’ Disease outbreaks.^14^ However the lack of standardized guidelines and resource limitations for building managers to implement WMP can lead to inadequate or inconsistent monitoring.^14^ Preventative controls that can be included in a WMP include hot water temperature regulation, nutrient limitation, and prevention of aerosolization to prevent *Legionella* exposure and growth in water systems.^5, 11^ Building level water treatment (physical, thermal and chemical) is typically deployed as an emergency measure in water systems that are highly colonized with *Legionella* and need immediate corrective action.^12^ Flushing is the most frequently used treatment for *Legionella* in buildings, where water lines are flushed to replace aged water with “fresh” water from the drinking water distribution system (DWDS).^15^ However, the long-term success of flushing for *Legionella* remediation is inconsistent across premise plumbing systems^16–19^. Thermal heat shocks are another widely used approach to eradicate *Legionella* in hot water systems after an outbreak situation.^12^ This approach requires heating hot water tanks to above 60 °C (140 °F) then circulating the heated water throughout hot-water lines to kill *Legionella*. This treatment typically achieves only short-term success and recolonization can occur within weeks to months after implementation.^12, 20^ Chemical shock treatments have been used for a more aggressive approach to treat *Legionella* in buildings by increasing the disinfectant (e.g. chlorine dioxide) in the water lines for bacterial eradication.^12, 21^ Hyperchlorination is a chorine-based disinfection at high concentrations (e.g. 10-50 mg/L Cl_2_) that inactivates pathogens in drinking water, but its effectiveness is dependent on pH and temperature in water.^22^ Chlorine dioxide and monochloramines are highly effective chlorine-based treatments for *Legionella* in potable water systems due to their efficacy at higher temperature and at wide range of pH.^22^ While treatments at the building level can reduce *Legionella* within the short-term (days to weeks), *Legionella* can survive within biofilm and resurge in drinking water from biofilm detachment weeks to months after treatment.^23^ Biofilms are difficult to treat and remove from premise plumbing systems, and as pipes age, more biofilm can accumulate with further sequestration of pathogens like *Legionella*.^24^

During the COVID-19 pandemic in the US and worldwide, many buildings experienced complete or partial shutdowns. These shutdowns reduced water use and increased the likelihood for *Legionella* growth within building water systems. These shutdowns, intended to prevent the spread of the COVID-19 infections, led to prolonged water stagnation and negative impacts on building water quality.^25–28^ The impact of decreased water demand on *Legionella* proliferation in non-healthcare settings has been shown to have a wide range of outcomes with from no detection to widespread detection of *L. pneumophila* during shutdowns.^19, 26, 28, 29^

Flushing is often recommended for the safe reopening of buildings after extended periods of stagnation.^13, 15, 30^. However, flushing has been shown to have either a short-term benefit to reduce *Legionella* in drinking water or led to an increase of *Legionella* occurrence in recommissioned buildings.^16, 17, 19, 31, 32^ Rhoads et al. found that water stagnation alone does not always result in *Legionella* growth, but reduced water demand in conjunction with other premise plumbing characteristics (i.e. hot-water recirculation, water-use patterns at individual outlets, external disturbances) contribute to increased *Legionella* occurrence in buildings.^31^ There is a need to evaluate building-level treatments beyond flushing during and after the COVID-19 building shutdown. Additionally, there are limited studies that monitor for *Legionella* in non-healthcare buildings beyond the several months from the start of the COVID-19 shutdown.^31^

The objectives of this study were to: (1) assess the water quality of hot-water lines in an aging residential building during and following the COVID-19 building shutdown over a two-year period (2) evaluate multiple water treatment approaches that were targeted towards reducing *L. pneumophila* colonization during intermittent water-use and occupancy. Physiochemical water quality parameters were also measured, and cold-water line samples were collected to assess *L. pneumophila* in drinking water from the entire building and the DWDS. This long-term study represents one of the few studies that extensively monitored for *Legionella* growth in a non-healthcare premise plumbing system during and long after the COVID-19 building shutdown. Results from this research provides insight on *Legionella* occurrence during different periods of building occupancy and the factors contributing to *Legionella* growth in real-time while building managers worked to reduce the risk of *Legionella* exposure and infection.

## MATERIALS AND METHODS

### Site Selection

Treatment interventions occurred in a residential building with known colonization of *Legionella* on an educational campus in Maryland, USA. This building, denoted Building A, experienced limited water-use with no occupancy from March 2020 to January 2021 due to the COVID-19 pandemic building shutdown implemented in March 2020. From January 2021 to May 2021, the building was partially re-occupied and full occupancy was initiated in September 2021. Building A was built in the mid-1950s and is comprised of four floors (Basement, Floor 1, Floor 2, Floor 3) and three wings i.e., risers (Riser A, Riser B, Riser C). A 4,000 L water heater tank located in the Basement of Riser B heats and stores the building’s hot water. The hot water was distributed in a recirculating system from the top to bottom floors (Figure 1). Twelve showers were selected for hot water sampling with at least three showers selected for Floors 1-3 and each riser. Cold-water was sampled from the basement (main line entry from the DWDS located in the Basement in Riser C) and two sink faucets that were most proximal and distal from the main line entry. The building’s potable water was supplied by the metropolitan drinking water utility which treats surface water for municipal use and distributes the water using chlorine for disinfectant residual throughout an urban environment.

**Figure 1.**
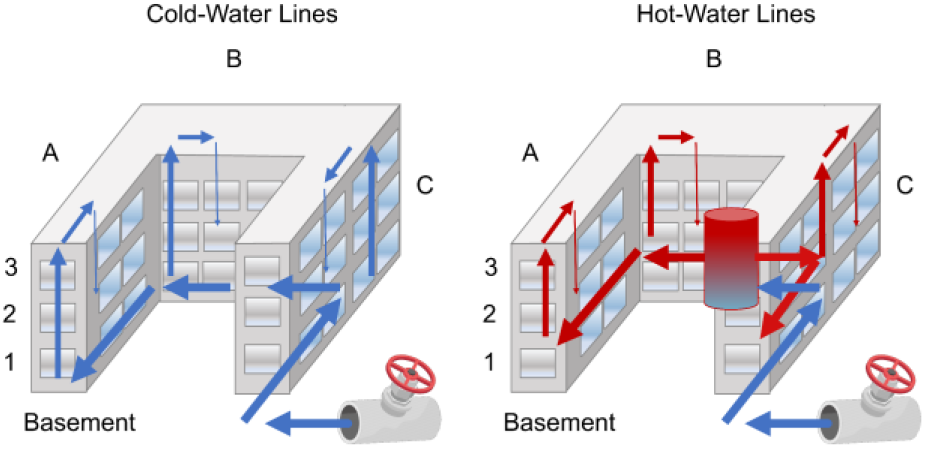
Building hydraulic flow of cold-water entering from the Drinking Water Distribution System (DWDS), hot-water heating, and flow. (Left) The cold-water from the DWDS enters through Riser C in the Basement and flows into Riser B and then Riser A, the cold-water then rises to the top 3rd Floor and descends to the 2nd and 1st Floors. (Right) The hot-water is heated first in a 4,000 L heat tank located in Riser B in the Basement. After heating, the hot-water flows to Risers A & C simultaneously, then rises to the 3rd Floor and descends to the 2nd and 1st Floors. The hot-water recirculates throughout the building’s main hot-water lines.

### Sample Collection

One-liter potable water samples were collected over the course of 22 months (August 2020 to May 2022) from Building A. Samples were collected in sterile polypropylene copolymer (PPCO) bottles from hot water lines at each of the 12 showers with shower heads removed, and cold-water lines at the entry point in the basement (“Inlet”) and two sink faucets with the aerators removed. First-draw samples, representing water at the outlet, were collected at time zero (T0) at every sampling point. A second sample was collected after a five-minute flush (T5), which represented water within the building pipes. Immediately after collection the samples were portioned into 100 mL in Whirl-Pak bags containing vessels with 25 mg sodium thiosulfate (Whirl-Pak; Fort Atkinson, Wisconsin) for *L. pneumophila* culture, and 40 mL portioned into amber glass vials for turbidity and total organic carbon (TOC) analysis. Aliquots were placed on ice and transported to the laboratory within two hours of collection.

### Culture of L. pneumophila

Culturable *L. pneumophil*a serogroup 1 to15 were processed and quantified using the IDEXX Legiolert reagents and protocol (IDEXX Laboratories; Westbrook, Maine). Undilute and ten-fold diluted water samples (100 mL) were processed on the same day of collection using the manufacturer’s potable water protocol or modified protocol (described in the Materials and Methods section of Supporting Information).^33^ For ten-fold dilutions, 10 mL of undiluted sample was diluted in 90 mL of 0.1% peptone solution per the manufacturer’s recommendation. After seven days of incubation at 39 °C and 85% humidity in a humidified incubation chamber, positive wells were enumerated, and the most probable number (MPN) calculated using the MPN table provided by IDEXX.

### Water Quality Analysis

Free chlorine, temperature, and pH were measured on-site immediately after collection to understand the factors associated with *Legionella* concentrations. Temperature and pH were measured using HACH HQ40d portable meter with pH meter probe PHC20101 (HACH; Loveland, Colorado). Free chlorine (mg/L Cl_2_) was measured using the DR300 Pocket Colorimeter (HACH; Loveland, Colorado). Turbidity was measured using an OAKTON Turbidity Meter (Cole-Parmer; Vernon Hills, Illinois) and TOC was analyzed using Sievers M9 TOC Analyzer (GE Analytical Instruments; Boulder, Colorado). All water quality analyses were conducted using respective manufacturer instructions.

### Potable Water Building Treatments

The following water treatment interventions were examined to understand their impact on reduction of *Legionella* bacteria from the potable water system.

#### Heat shock

Single heat shock treatments were conducted on August 9, 2020 (Heat Shock 1), December 12, 2020 (Heat Shock 2), and May 18, 2021 (Heat Shock 3). The hot water pipe temperature was elevated to 65 °C by heating the 4,000 L hot water tank located in the Basement of Riser B. The hot water in the hot water pipes was flushed from all outlets until the targeted temperature (65 °C) was measured. After the elevated hot water rested in the lines for 12 hours, the hot water tank temperature was reduced to the operational temperature of 49 °C. The hot-water was flushed again from every outlet until the normal temperatures (49 °C) was obtained.

#### Chemical shock

Single chemical shock treatments using chlorine dioxide or chlorine were conducted on July 15, 2021 (Chlorine Dioxide 1), August 10, 2021 (Chlorine Dioxide 2), and August 16, 2021 (Hyper-Chlorination). For the single chlorine dioxide shocks, the hot and cold-water lines were treated with 2.5 mg/L of chlorine dioxide (ClO_2_) by injecting disinfectant at the diversion of hot and cold-water pipelines in the basement. Water was flushed at every outlet from both hot and cold-water lines until the targeted 2.5 mg/L ClO_2_ was reached. After resting for 12 hours of treatment, water from both water lines were flushed from outlets until a residual of 1.0 mg/L ClO_2_ was measured. For the single Hyper-Chlorination shock, hot and cold-water lines were treated by injection of a high concentration of chlorine (Cl_2_) at 20 mg/L. Similar to the chlorine dioxide treatment, both water lines were flushed to reach the targeted chlorine concentration, rested in the lines for 12 hours, then were flushed from outlets until free chlorine residuals were below 4 mg/L Cl_2_. All single thermal and chemical shock treatments were conducted when the building was vacant to ensure safety of building occupants.

#### Continuous low-level treatment protocol

An on-premise continuous low-level chlorine dioxide was implemented on February 21, 2022, and continued long-term (10 weeks later). Both hot and cold-water lines were treated by injecting 0.2-0.4 mg/L ClO_2_ of stable chlorine dioxide into the water main prior to the diversion into hot and cold-water pipelines.

### Data Analysis

Statistical tests and data transformations were performed with STATA (Stata 10 Data Analysis and Statistical Software; StataCorp LP, College Station, Texas). On non-normally distributed *L. pneumophila* data, two-sided Wilcoxon Signed Rank Test were performed on paired samples between sampling events/treatment interventions and between first draws (T0) and five-minute flush (T5). Non-parametric Kruskal Wallis H hypothesis test was performed to compare samples between variables with multiple building characteristics (i.e., Risers, Showers). Correlation between *L. pneumophila* and physiochemical parameters were assessed with Kendall Tau Coefficient Correlation test with log-transformed *L. pneumophila* concentrations below the limit of detection (<1 MPN/100mL) substituted as 0 (i.e., log_10_(1)). Positive samples were indicated by *L. pneumophila* results with ≥1 MPN/100 mL. Statistical significance was defined a priori using p=0.05. All graphs were produced by Microsoft Excel and STATA software.

## RESULTS

### Microbiological Analysis

A total of 624 samples were collected from hot-water lines with an additional 121 samples collected from cold-water lines (total N=745). All samples were tested and quantified for culturable *L. pneumophila* Serogroup 1 to 15. Culturable *L. pneumophila* were detected in all hot-water shower locations and at both times of collection (T0 and T5) with results ranging from <1 MPN/100mL to >22,736 MPN/100mL (Figure 2). The overall percent positivity for *L. pneumophila* in all hot water samples was 76.6% (n=478/624) with a geometric mean of 139 MPN/100 mL. Hot-water samples collected at T0 had a geometric mean of 245 MPN/100mL with 73.0% positivity (n=219/300) and those at Time 5 had a geometric mean of 83 MPN/100mL with 78.3% positivity (n=235/300). Statistical analysis found a significant 0.47 log decrease of *L. pneumophila* concentrations from T0 to T5 samples across matched samples (p<0.001).

**Figure 2.**
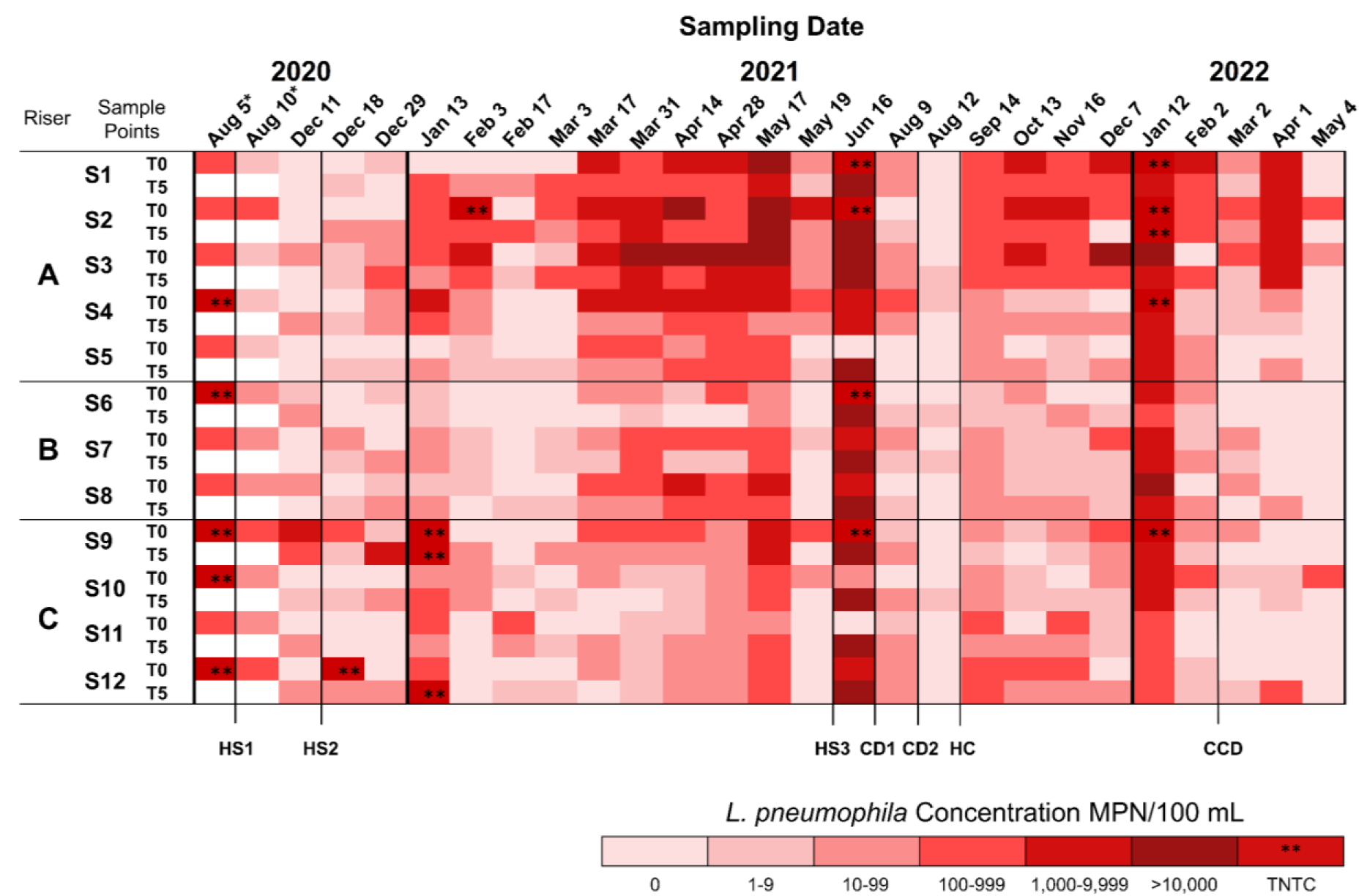
Heatmap of culturable *L. pneumophila* in Building A premise plumbing system from hot-water lines from August 5, 2020 to May 4, 2022. Values shown on heatmap were the higher concentration results between undilute and ten-fold dilution samples. S1-12 represent Shower 1 through Shower 12. T0 represents results at first draw and T5 after a five-minute flush. *Samples collected on August 5, 2020 & August 10, 2020 were collected only after a one-minute flush. **TNTC values that were too numerous to count (>22,736 MPN/100 mL). HS: Heat Shock. CD: Chlorine Dioxide. HC: Hyper-Chlorination. CCD: Continuous Chlorine Dioxide. White space indicates no collection of sample.

Riser A had the highest statistically significant concentration of *L. pneumophila* with a geometric mean (GM) of 312 MPN/100 mL and 80.8% positivity (n=210/260) compared to Riser B with a GM of 62 MPN/100 mL with 75.0% positivity (n=117/156) and Riser C with a GM of 84 MPN/100 mL with 72.6% positivity (n=151/208) (p<0.001) (Figure S1). Shower 2 (Riser A) had the highest statistically significant GM of *L. pneumophila* (1,114 MPN/100 mL; p<0.001) and Shower 3 (Riser A) had the highest statistically significant positivity (92.3% ; p=0.007) (Figure S1). Shower 6 (Riser B) had the lowest statistically significant GM of *L. pneumophila* (41 MPN/100 mL; p<0.001) and Shower 11(Riser C) had the lowest statistically significant positivity (59.6%; p=0.007) (Figure S1).

The overall percent positivity for cold-water samples was 42.2% (n=51/121) with a GM of 46 MPN/100 mL. The majority of cold-water samples at the Inlet were negative for *L. pneumophila* (GM: 2 MPN/100 mL; 2.2% positivity; n=45) (Figure S2). The distal cold-water location experienced a higher GM of *L. pneumophila* (59 MPN/100 mL; 73.3% positivity; n=30) compared to proximal location (43 MPN/100 mL;60.9% positivity; n=46) (Figure S2). Cold-water samples from sink locations collected at T0 had a GM of 115 MPN/100mL with 68.4% positivity and at T5 had a GM of 20 MPN/100mL with 63.1% positivity. Across matched cold-water samples, there was a statistically significant 0.76 log decrease of *L. pneumophila* concentrations from T0 to T5 (p<0.001).

### Building Water Treatment Effects

#### Single Heat Shock Treatments

Samples prior to and after each water treatment were compared using paired statistical analysis. The hot-water samples after Heat Shock 1 experienced a significant 1.50 log reduction of culturable *L. pneumophila* concentration (p=0.005) between samples collected four days prior (August 5, 2020) and one day after treatment (August 10, 2020) (Figure 3A). *L. pneumophila* concentrations remained constant afterwards four months later (December 11, 2020) (p=0.21). The analysis for Heat Shock 2 did not find a significant reduction in *L. pneumophila* (p=0.87) between samples collected one day prior (December 11, 2020) and six days after treatment (December 18, 2020) (Figure 3B). There was a significant 0.20 log increase of *L. pneumophila* comparing samples prior to Heat Shock 2 (December 11, 2020) and two weeks after treatment (December 29, 2020) (p=0.05) (Figure 3B). For Heat Shock 3, samples collected one day prior (May 17, 2021) and one day after treatment (May 19, 2021) experienced a significant 1.53 log reduction of *L. pneumophila (*p<0.001), but comparison of pre-Heat Shock 3 samples to those four weeks after treatment (June 16, 2021) found no significant difference in *L. pneumophila* concentrations (p=0.92) (Figure 3C).

**Figure 3.**
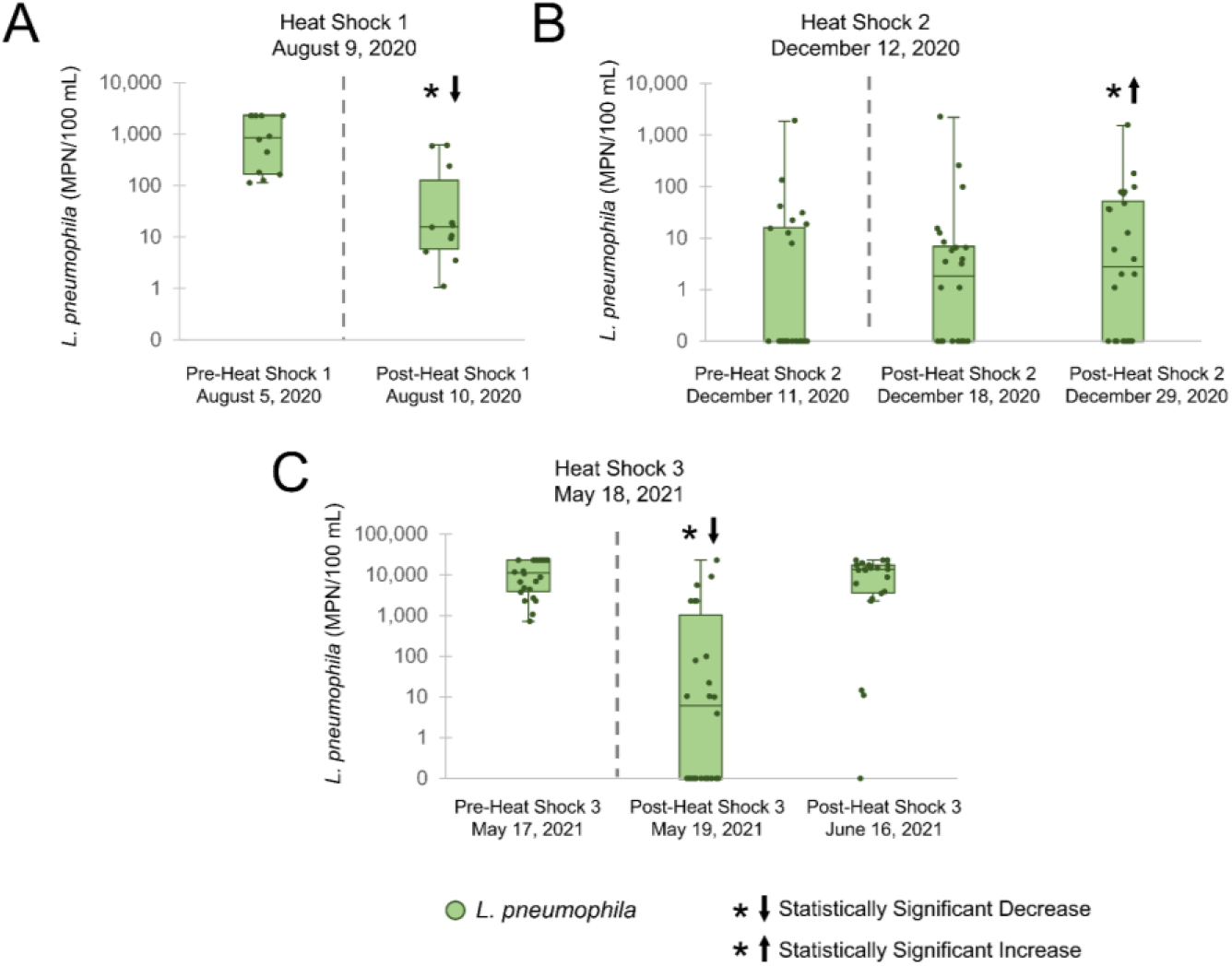
Single heat shock treatments conducted in Building A. Hot-water samples were compared using Wilcoxon Signed Rank Test on paired *L. pneumophila* shower results. (A) Samples (n=12) were compared for reduction of *L. pneumophila* four days prior (Pre-Heat Shock 1) to one day after (Post-Heat Shock 1) treatment conducted on August 9, 2020 (Heat Shock 1). (B) Samples (n=24) were compared for reduction on *L. pneumophila* one day prior (Pre-Heat Shock 2) to six days after (Post-Heat Shock 2) and two weeks after (Post-Heat Shock 2) treatment conducted on December 12, 2020 (Heat Shock 2). (C) Samples (n=24) were compared for reduction of *L. pneumophila* one day prior (Pre-Heat Shock 3) to one day after (Post-Heat Shock 3) and four weeks after (Post-Heat Shock 3) treatment conducted on May 18, 2021 (Heat Shock 3). *↓ There was a statistically significant decrease in *L. pneumophila* compared to samples prior to treatment. *↑There was a statistically significant increase *L. pneumophila* compared to samples prior to treatment.

#### Single Chemical Shock Treatments

Chemical shock treatments were conducted and evaluated after the multiple single Heat Shock treatments failed to provide long-term reduction for *L. pneumophila*. Analysis of Chlorine Dioxide 1 samples collected four weeks prior (June 16, 2021) to four weeks after (August 9, 2021) treatment found a significant 2.29 log reduction of *L. pneumophila* (p<0.001) (Figure 4A). Results for Chlorine Dioxide 2 found a significant 1.29 log reduction of *L. pneumophila* (p<0.001) between samples collected one day prior (August 9, 2021) and two days after treatment (August 12, 2021) (Figure 4B). Comparison of samples collected four days prior to Hyper-Chlorination (August 12, 2021) and four weeks after (September 14, 2021) found a significant 1.80 log increase in *L. pneumophila* (p<0.001) (Figure 4C).

**Figure 4.**
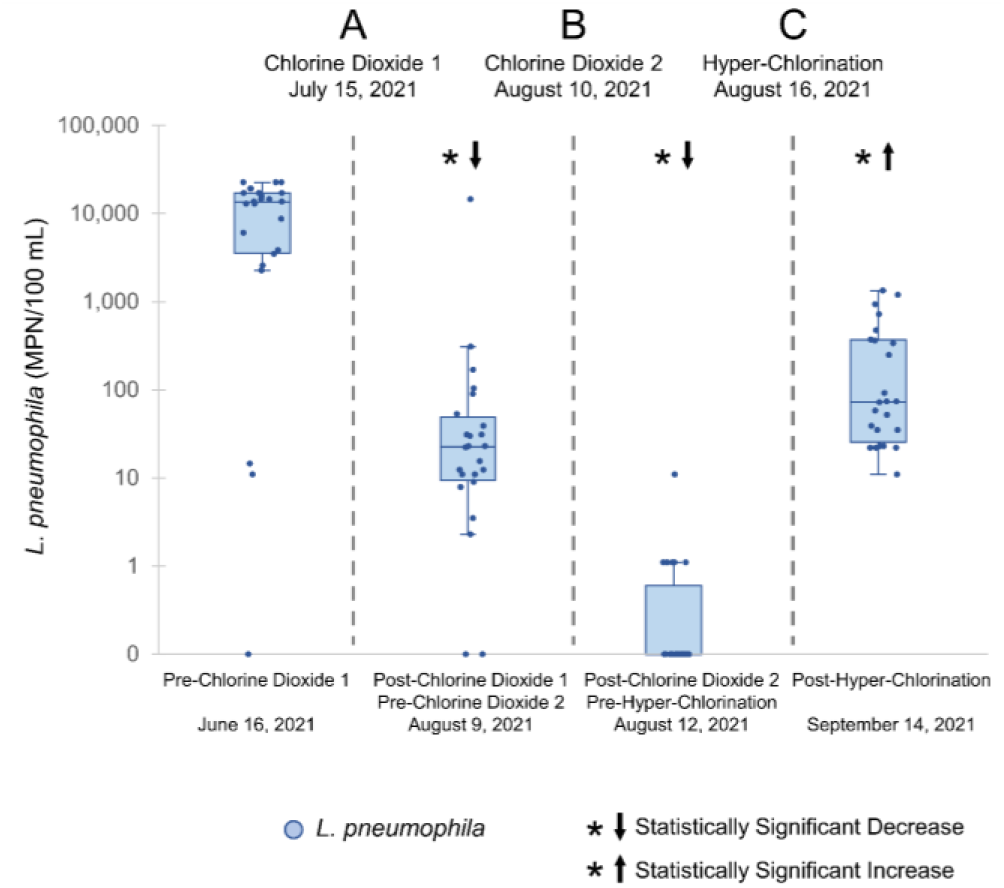
Single chemical shock treatments conducted in Building A. Hot-water samples were compared using Wilcoxon Signed Rank Test on paired *L. pneumophila* shower results. (A) Samples (n=24) were compared for reduction on *L. pneumophila* four weeks prior (Pre-Chlorine Dioxide 1) to three weeks after (Post-Chlorine Dioxide 1) treatment conducted on July 15, 2021 (Chlorine Dioxide 1). (B) Samples (n=24) were compared for reduction of *L. pneumophila* one day prior (Pre-Chlorine Dioxide 2) to two days after (Post-Chlorine Dioxide 2) treatment conducted on August 10, 2021(Chlorine Dioxide 2). (C) Samples (n=24) were compared for reduction of *L. pneumophila* four days prior (Pre-Hyper-Chlorination) to four weeks after (Post-Hyper-Chlorination) treatment conducted on August 16, 2021 (Hyper-Chlorination). *↓ There was a statistically significant decrease in *L. pneumophila* compared to samples prior to treatment.*↑ There was a statistically significant increase *L. pneumophila* compared to samples prior to treatment.

#### Continuous Low-Level Chlorine Dioxide Treatment

Single chemical shock treatments were unable to control for *L. pneumophila* long-term (Figure 4), therefore a continuous low-level chemical treatment was evaluated. Comparison of samples prior to implementation of treatment (February 2, 2022) and one week later (March 2, 2022) found a significant 0.58 log reduction in *L. pneumophila* (p<0.001) (Figure 5). Six weeks after implementation (April 1, 2022), *L. pneumophila* concentrations returned to pre-implementation levels (p=0.13). Ten weeks after implementation (May 4, 2022), there was a significant 0.79 log reduction in *L. pneumophila* (p<0.001) (Figure 5).

**Figure 5.**
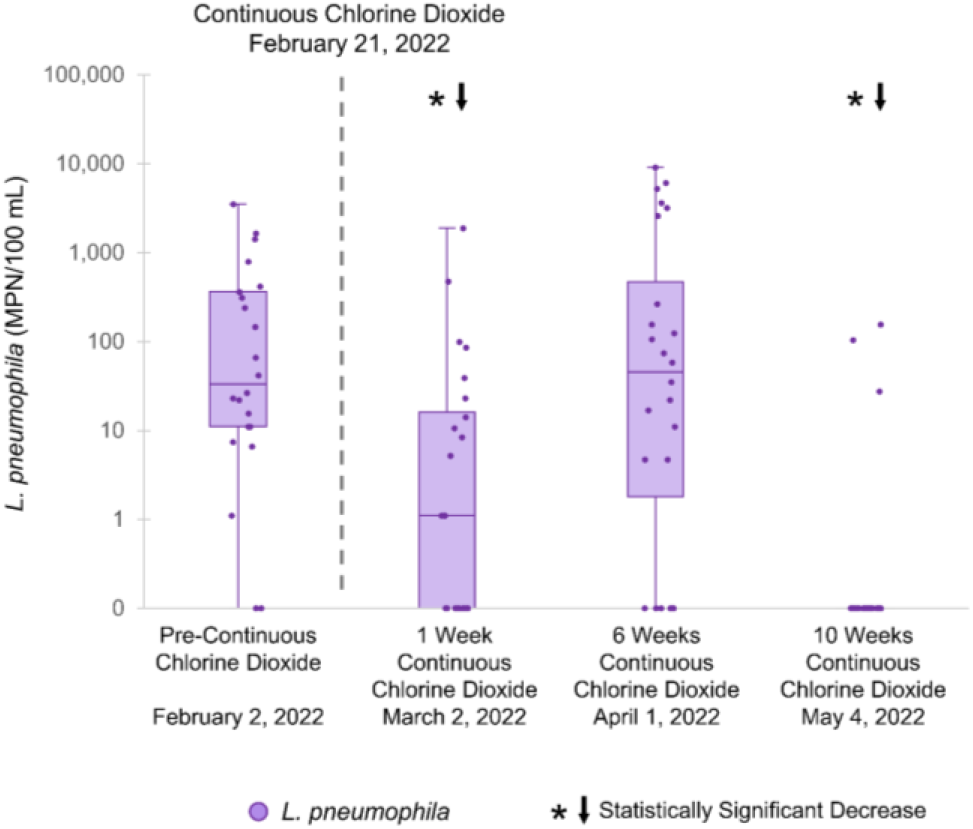
Continuous low-level chlorine dioxide treatment implemented in Building A. Hot-water samples were compared using Wilcoxon Signed Rank Test on paired *L. pneumophila* shower results (n=24). Samples were compared for reduction of *L. pneumophila* three weeks prior (Pre-Continuous Chlorine Dioxide) to one week, six weeks, and ten weeks after treatment implemented on February 21, 2022 (Continuous Chlorine Dioxide). *↓ There was a statistically significant decrease in *L. pneumophila* compared to Pre-Continuous Chlorine Dioxide samples.

### Water Quality Indicators

#### Free Chlorine

Residual free chlorine concentrations in cold-water samples ranged from below the limit of detection (<0.02 mg/L Cl_2_) to 1.06 mg/L Cl_2_. The Inlet cold-water sample from the DWDS had a mean concentration of 0.49 mg/L Cl_2_ (Range: <0.02 to 0.82 mg/L Cl_2_) (Figure S3). Chlorine levels in the proximal cold-water samples averaged 0.24 mg/L Cl_2_ (Range: <0.02 to 1.06 mg/L Cl_2_) compared to chlorine in the distal cold-water samples of 0.13 mg/L Cl_2_ (Range: <0.02 to 0.67 mg/L Cl_2_) (Figure S3). Due to loss of chlorine at elevated temperatures, hot-water samples had low concentrations on average (Mean: <0.1 mg/L Cl_2_; Range: <0.02 to 0.97 mg/L Cl_2_) at each riser and shower (Except Shower 2: mean 0.13 mg/L Cl_2_) (Figure S3). At each cold-water sink location, residual free chlorine did statistically increase across matched samples from T0 to T5 (T0 mean: 0.25 mg/L Cl_2_; T5 mean: 0.36 mg/L Cl_2_) (p<0.001). After Building A was re-occupied with partial water-use (January 2021), chlorine residual levels (hot and cold-water samples) significantly increased after three weeks (p=0.01) but did not increase further after full re-occupancy (September 2021) with resumed normal water-use (p=0.76). Within cold-water lines, free chlorine had a moderate negative correlation with *L. pneumophila* concentrations (Kendall tau-b= −0.47, p<0.001) indicating that samples with higher free chlorine residuals resulted in reduced *L. pneumophila* concentrations. Samples had a 70.7% (n=58) positivity for *L. pneumophila* when free chlorine concentrations were less than or equal to 0.2 mg/L Cl_2_ and 12.7% (n=63) positivity when concentrations were greater than 0.2 mg/L Cl_2_.

#### Temperature

Hot-water temperatures were significantly different between T0 and T5 (p<0.001). T0 hot-water samples averaged lower temperatures (Mean: 26.1 °C; Range: 15.6-37.4 °C) compared to T5 hot-water samples (Mean: 38.0 °C; Range: 22.1-50.9 °C) (Figure S4). The Inlet cold-water samples had the lowest measured temperatures (Mean:14.9 °C; Range: 8.7-22.5 °C) while the proximal and distal had higher cold-water temperatures (Mean: 23.1 °C; Range: 12.6-33.1 °C).

*L. pneumophila* growth occurred in samples with temperatures ranging from 14.6 °C to 49.8 °C, with the majority of the positive *L. pneumophila* samples (70.4%) at temperatures within the range suitable for *Legionella* growth (20 °C to 45 °C).^34–36^ Negative *L. pneumophila* samples occurred in temperatures ranging from 8.7 °C to 50.9° C. Correlation analysis found no association between hot-water line temperatures and *L. pneumophila* concentrations (Kendall tau-b= −0.005, p=0.85). There was a moderate positive association between cold-water line temperatures and *L. pneumophila* concentrations (Kendall tau-b=0.50, p<0.001).

#### pH

The pH ranged from 6.5 to 8.6 (Mean: 7.4) in both hot and cold-water samples. Positive samples occurred at pH range of 6.5-8.4. Among hot-water samples, there was a significant increase in pH between T0 and T5 samples (p<0.001), with average T0 pH (Mean=7.2) slightly lower than the average T5 pH (Mean=7.50) (Figure S5). The cold-water samples also experienced a significant increase in average pH between T0 (pH=7.25) and T5 (pH=7.50) samples (p<0.001) (Figure S5). A strong negative correlation was found between *L. pneumophila* and cold-water samples with a pH higher than 7.9 (Kendall tau-b= −0.62, p=0.004).

## DISCUSSION

There was high colonization found in the hot water system of Building A and the treatment found to be most effective for long term reduction of *Legionella pneumophila* was continuous low-level chlorine dioxide dosing. While single heat shock and chemical shock treatments reduced *L. pneumophila* in the short-term, these single treatments were ineffective in controlling *L. pneumophila* in the building long-term. Single shock treatments may not be efficient in complete removal of biofilm that can sequester *L. pneumophila* and surviving *L. pneumophila* can repopulate the water system after treatment.^37, 38^ Continuous low-level chlorine dioxide treatment was effective in reducing *L. pneumophila* in hot water lines long-term.

### Building Characteristics and Water Quality Indicators for *L. pneumophila*

Similar to other studies, *L. pneumophila* was present in both hot-water lines and cold-water lines. Hot-water lines experienced higher positivity and concentration in Riser A which may be indicative of higher colonization of *L. pneumophila* in Riser A pipes compared to Riser B and C. The difference in colonization by riser may be explained by the circulation of hot-water in the building. Building A is designed with a recirculating hot-water system with a storage tank. Hot water storage tanks have been linked to increased *Legionella* infection risk due to temperature ranges amenable to *L. pneumophila* growth (20-45 °C) and stagnant water conditions.^39, 40^ Additionally, suboptimal recirculating systems, with temperatures below 48 °C to prevent scalding, can favor *Legionella* growth in parts of the hot-water circulation system.^40, 41^ Flushing showers for five minutes reduced *L. pneumophila* concentrations across all hot-water samples (T0 vs T5) from individual outlets (p<0.001). While frequently recommended, short-term flushing of hot-water systems, particularly in recirculating hot-water systems, may not be effective in *Legionella* reduction long-term since *Legionella* can recirculate and return to shower outlets after flushes (Figure 2).^11,40^ The cold-water samples experienced high *L. pneumophila* positivity in the proximal and distal locations, but minimal positivity at the Inlet, which may indicate colonization of *L. pneumophila* in the cold-water pipes and minimal influence from the DWDS (Figure S2). While *Legionella* prefer warmer temperatures, these bacteria can still survive and grow within cold-water systems.^42^ Compared to the hot-water lines, the cold-water samples had lower *L. pneumophila* positivity and concentrations, which may be due to higher retention of free chlorine residual and temperatures below *Legionella’s* optimal growth range.^11, 43^

The proximal location sample sites experienced lower *L. pneumophila* levels compared to distal locations which may be correlated to the reduction of free chlorine residual as it travels through the building premise plumbing system to distal sites (Figure S3).^44^ Studies have found a higher free chlorine residual was associated with lower *Legionella* growth, but as conditions become less optimal for free chlorine (increased pH and higher temperature), the residual becomes less effective in *Legionella* reduction.^45^ In addition to the impact of pH on free chlorine disinfection effectiveness, pH has also been known to directly reduce *Legionella* growth at higher pH (8.2-10.5 pH).^46, 47^ Although our sample size was limited, cold-water samples with higher pH (>7.9) were associated with lower *L. pneumophila* concentrations despite lower chlorine residual in these samples (Range: <0.02-0.46 mg/l Cl_2;_ n=14).

### Prolonged Stagnation and Building Treatments

Building A experienced extended stagnation for nine months from March 2020 to December 2020 and in response, building managers implemented interventions in the hot and cold-water lines in accordance with the CDC’s published guidance to reduce *Legionella.*^11, 30, 48^ Heat Shock 1 and Heat Shock 2 resulted in a reduction *L. pneumophila* short-term, however prolonged water stagnation may have exacerbated growth weeks later (Figure 3A and Figure 3B).^49^ The building was partially re-occupied on January 13, 2021, after the shutdown, and with the return of partial water-use in the building, *L. pneumophila* concentrations substantially rose months later (Figure 2). Heat Shock 3 resulted in a short-term reduction of *L. pneumophila* then a rapid return to pre-heat shock levels in majority of sampling sites in the building (Figure 3C). Guidance from the American Water Works Association (AWWA) does not recommend heat shock treatments due to the rapid bounce back of *Legionella* concentrations.^50^ The heat resilience within biofilms may have also contributed to the *L. pneumophila* resurgence after heat shock treatments.^50^ After the single heat shocks failed to control *L. pneumophila*, building operators turned to chemical disinfection to treat the hot and cold-water systems. The first chlorine dioxide treatment (Chlorine Dioxide 1) was effective in reducing *L. pneumophila* and the subsequent chlorine dioxide treatment (Chlorine Dioxide 2) further temporarily ablated *L. pneumophila* to levels below detection (Figure 4A and 4B). An additional chlorination shock (Hyper-chlorination) was conducted to further eradicate *L. pneumophila* from the system but experienced an increase of *L. pneumophila* weeks later (Figure 4C). A possible explanation for *L. pneumophila* resurgence could be the detachment of biofilm resulting from the chlorine shocks.^51^ As the building transitioned back to limited water-use from short term vacancy (December 7, 2021 to January 12, 2022), *L. pneumophila* concentrations increased further (Figure 2). The limited use may have resulted in water stagnation that may have contributed to further biofilm formation within the pipes.^52^ As a response to the failure of single chemical treatments on long-term *L. pneumophila* control, building managers shifted to evaluating a continuous low-level chemical treatment approach. This decision was made to provide continuous chemical treatment at levels that were safe for building occupants.^53^ After the implementation of the continuous low-level chlorine dioxide treatment, *L. pneumophila* concentrations stabilized and finally reduced months after the implementation of treatment (Figure 5). Since the levels of continuous chlorine dioxide treatments were low compared to shock treatments, it may have taken months longer to experience the full effect of the disinfection. Our findings are similar to several studies that have also shown the long-term effectiveness of continuous chlorine dioxide treatment in potable water systems.^54, 55^

Examining the cold-water lines in this building was necessary to determine potential *Legionella* exposure from the entire premise plumbing system. It should be noted that when a continuous treatment approach was selected, building managers applied for and received supplemental disinfection permits for building level treatment. One limitation of the study was the consistency of sampling events and sample collection. The remediation challenges in Building A were addressed in real-time, therefore there were constraints on consistent sampling. While the researchers were prepared to collect samples on a periodic basis, the scheduling of collection was constrained by operational decisions of the building managers.

### Future Directions of Water Management Programs

Multiple organizations including American Society of Heating, Refrigerating and Air-Conditioning Engineers (ASHRAE), American Industrial Hygiene Association (AIHA), AWWA, and CDC have published guidance for designing WMPs targeted at preventing the growth and spread of *Legionella* and other opportunistic pathogens in premise plumbing water systems.^11, 41, 50, 56^ Despite the recommendation for formal WMPs across potable water systems, very few buildings have a developed WMP due to limited knowledge and resources.^28^ Even among WMP that are well-developed, WMP can be poorly implemented which can be due to lack of expertise and long-term investment for *Legionella* control.^28^ In many instances, these guidance have recommend flushing to reduce *Legionella* from potable water systems in premise plumbing, however our study has demonstrated that flushing has minimal effect on *Legionella* control and more extensive treatment is required especially for a building highly colonized with *Legionella*.^11, 41^ Our results suggest continuous low-level treatment would be the best method for control of *Legionella* in highly colonized buildings. Continuous low-level treatment can provide sustained levels of disinfection that would control for *Legionella* growth, prevent the formation of biofilm, and remove existing biofilm over time.^57^ However considerations for cost and regulations for potable water treatment can be limiting factors for implementation.^58^

Furthermore, current published guidelines propose using chemical or physical water quality predictors be used to assess *Legionella* risk or control in premise plumbing systems.^11, 41^ Testing parameters such temperature, pH, and free chlorine have been recommended to predict the presence or absence of *Legionella* at exposure points (shower and sink outlets).^11, 59, 60^ Despite these suggestions, multiple studies have shown inconsistent relationships between surrogate parameters and *Legionella.*^11, 60, 61^ Overall, culture-based detection is essential for monitoring *Legionella* and should not be replaced by surrogates alone. Along with an improved sampling approach, there is a need for more evidence-based guidance in preventing *Legionella* within premise plumbing that can be widely accessible and inexpensively executed.^28, 61, 62^ Understanding a building’s characteristics and hydraulic engineering is critical for accurately monitoring for *Legionella* in a premise plumbing system. This study demonstrates that a well-developed and properly implemented WMP can reduce *Legionella* risk and prevent waterborne disease outbreaks.

## ASSOCIATED CONTENT

### Supporting Information

The following files are available free of charge. Supporting Information (PDF).

Experimental materials and methods, including sample collection and processing; supplemental results, including water quality parameters (turbidity and total organic carbon); figures showing *L. pneumophila* in hot-water samples, *L. pneumophila* in cold-water samples, mean free chlorine concentrations, mean temperatures, mean pH, mean turbidity, and mean total organic carbon concentrations.

### Notes

The authors declare no competing financial interest.

## Data Availability

All data produced in the present study are available upon reasonable request to the authors.

## ACKNOWLEDGMENT

The authors thank the building directors, managers, and operators for their assistance with building access and sampling. The authors also thank The Osprey Foundation of Maryland and the National Institute of Health, USA (NIH Grant T32 ES007141) for their financial support for this research.

