## Supplemental Information for "Two-Year Evaluation of *Legionella* in an Aging Residential Building: Assessment of Multiple Potable Water Remediation Approaches"

10 pages: supporting text, 7 supporting figures.

### SI Materials and Methods

#### Sample Collection and Processing

All samples were collected by team members who wore personal protective equipment (N95 masks and nylon gloves) to protect against inhalation exposure and prevent cross-contamination. At each shower or sink faucet location, the showerheads or aerators were removed prior to sampling. All shower valves were rotated to the maximum hot temperature designation to obtain only hot water at first-draw (T0) and five-minute flush (T5). Flow rates were calculated for each shower location and ranged from 5 L/min to 9 L/min (Median 5 L/min). Samples were inverted immediately for mixing after collection, and aliquoted into a 100 mL and Whirl-Pak vessel containing sodium thiosulfate (Whirl-Pak; Fort Atkinson, Wisconsin) for Legiolert assays. The remaining sample volume was used for physiochemical parameters with 30 mL for pH and temperature, 30 mL for turbidity, and 10 mL for TOC. Aliquots were stored at 4 °C in coolers and transported to laboratory immediately after collection.

Microbiological samples were brought up to room temperature before analysis and if needed, were adjusted for hardness per the Legiolert protocols using the Legiolert Supplemental Kit (IDEXX Laboratories; Westbrook Maine). For ten-fold dilutions, 10 mL of undilute sample was diluted into 90 mL of 0.1% peptone. Legiolert media (IDEXX Laboratories; Westbrook Maine) was then added to both undilute and dilute samples and shaken immediately. Samples were then placed into a 37 °C water-bath for 20 minutes to assist in complete dissolving of media. Subsequently, samples were poured into Legiolert Quanti-trays (IDEXX Laboratories; Westbrook, Maine) and face-up trays (to keep the back of the trays hydrated in each MPN pillow) were incubated at 39 °C and 85% humidity for 7 days. A field negative control consisting of 1 L of sterile MilliQ water transported to and from the sample site was processed and analyzed with each sample set.

### SI Results

#### Water Quality Parameters

*Turbidity.* All samples measured for turbidity were expressed in Nephelometric Turbidity Units (NTU) and ranged from non-detect (<1 NTU) to 174 NTU. Cold-water samples averaged higher turbidity (10.9 NTU; n=120) compared to hot-water samples (3.2 NTU; n=624). The Inlet cold-water samples experienced the highest average turbidity concentration (23.9 NTU) compared to the proximal (5.0 NTU), distal (2.4 NTU), and hot-water samples (Riser A: 3.2 NTU, Riser B: 3.2 NTU, Riser C: 3.4 NTU) (Figure S6). *L. pneumophila* positive samples were present in turbidity ranging from 1 to 78.8 NTU. Approximately 65.9% (n =743) positive samples had turbidity ≥ 1 NTU. Correlation analysis found a weak negative correlation between turbidity and *L. pneumophila* (Kendall tau-b=-0.11, $p<0.001$; n=744).

*Total Organic Carbon.* All samples were measured for total organic carbon (TOC) in parts per million (ppm). Hot-water samples experienced a wider range of TOC (Range: non-detect (<0.1)-10.9 ppm; Mean: 1.27 ppm) compared to cold-water samples (Range 0.23-6.83 ppm; Mean: 1.42). Only in the hot-water samples was there a significant difference of TOC concentration between T0 (Mean: 1.33 ppm) and T5 (Mean: 1.17 ppm), with average TOC slightly reduced at T5 ($p=0.01$) (Figure S7). The distal cold-water samples experienced slightly lower concentration of TOC (Mean: 1.26 ppm) compared to the Inlet (Mean: 1.47 ppm) and the proximal (Mean: 1.50 ppm) samples (Figure S7). *L. pneumophila* positive samples were present at TOC ranging from <0.1 to 10.9 ppm and had 65.9% (n=745) positivity with TOC>0. Correlation analysis found a weak negative association between *L. pneumophila* and TOC (Kendall tau-b=-0.22, p<0.001; n=743).

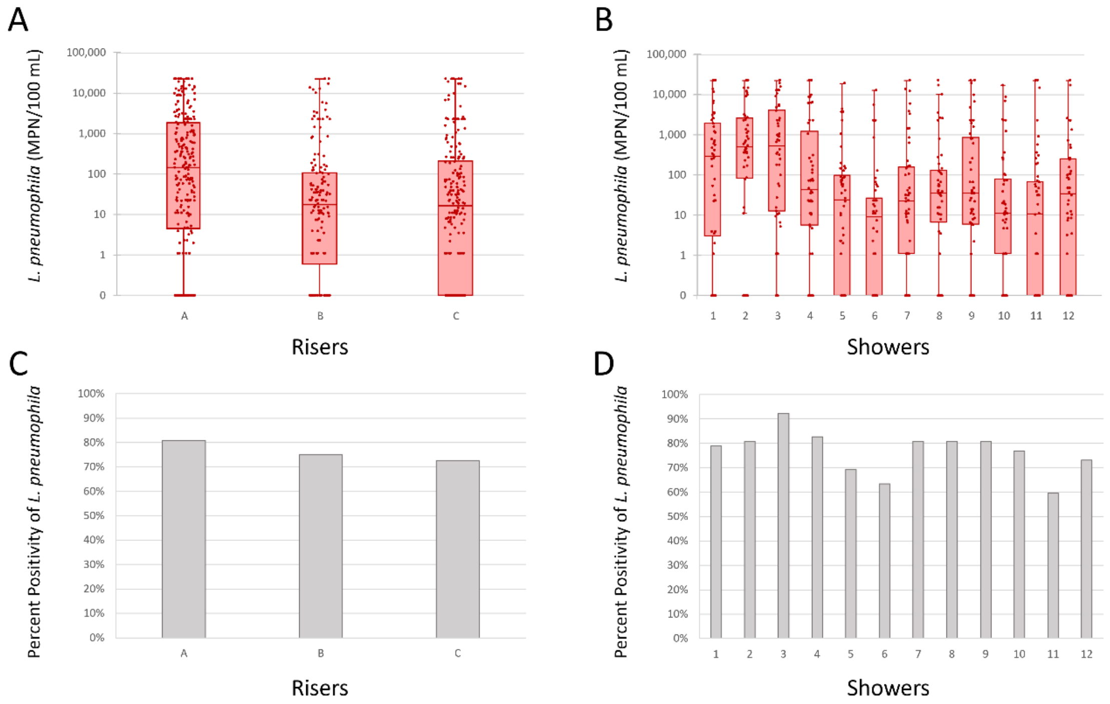


**Figure S1.** A) *L. pneumophila* concentrations across Riser A, B, and C hot-water samples. B) *L. pneumophila* concentrations across 12 showers from hot-water samples. C) Percent positivity of *L. pneumophila* concentrations across Riser A, B, and C from hot-water samples. D) Percent positivity of *L. pneumophila* concentrations across 12 showers from hot-water samples.

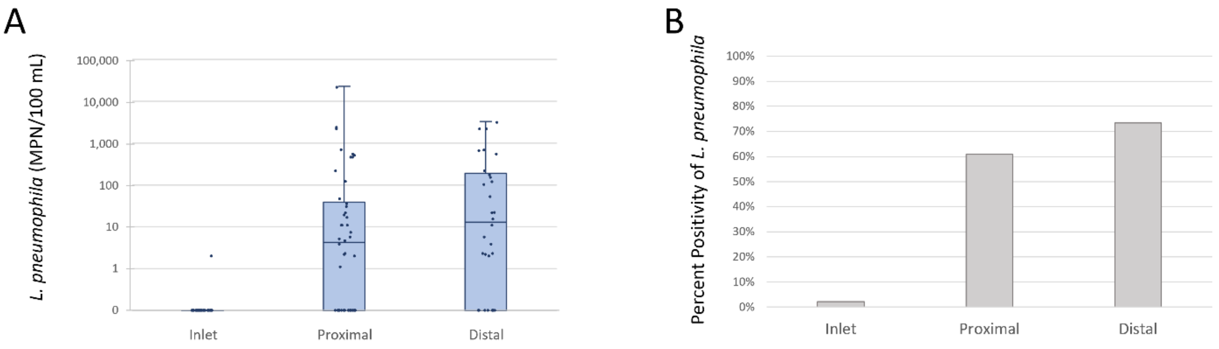


**Figure S2.** A) *L. pneumophila* concentrations across locations from cold-water samples. B) Percent positivity of *L. pneumophila* concentrations across locations from cold-water samples.

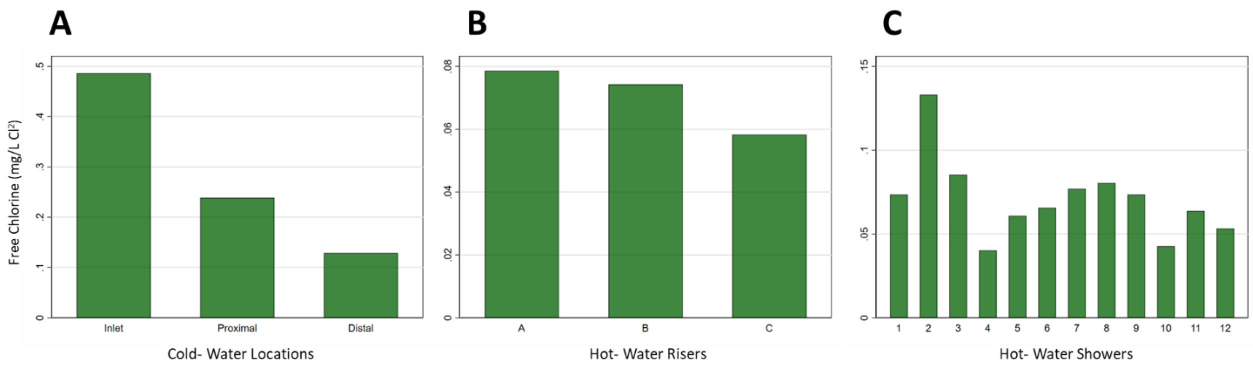


**Figure S3.** A) Free chlorine mean concentration across cold-water sample locations. B) Free chlorine concentration across risers from hot-water samples. C) Free chlorine concentration across 12 showers from hot-water samples.

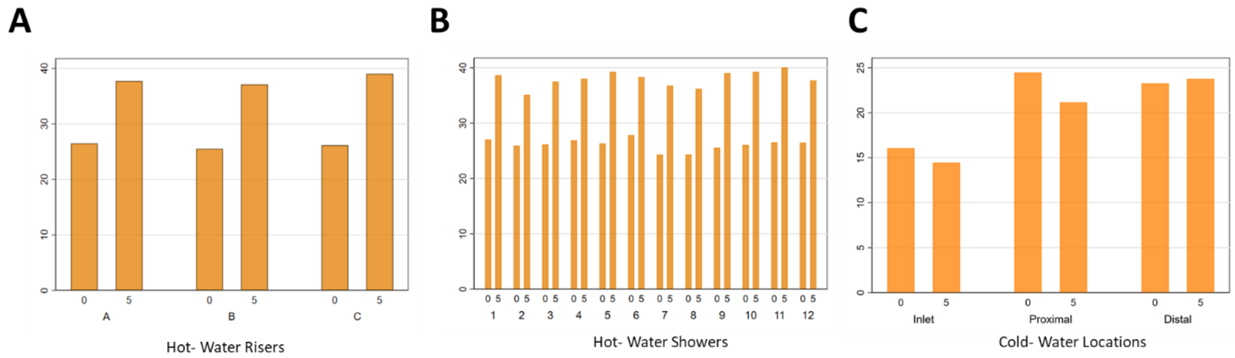


**Figure S4.** A) Mean temperatures across risers from hot-water samples. B) Mean temperatures across 12 showers from hot-water samples. C) Mean temperatures across cold-water sample locations. “0” indicates Time 0 and “5” indicates Time 5.

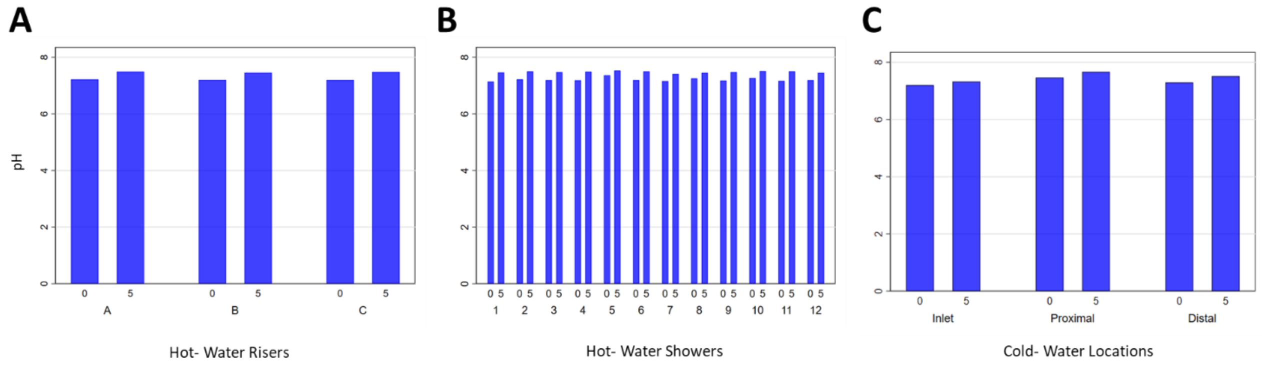


**Figure S5.** A) Mean pH across risers from hot-water samples. B) Mean pH across 12 showers from hot-water samples. C) Mean pH across cold-water sample locations. “0” indicates Time 0 and “5” indicates Time 5.

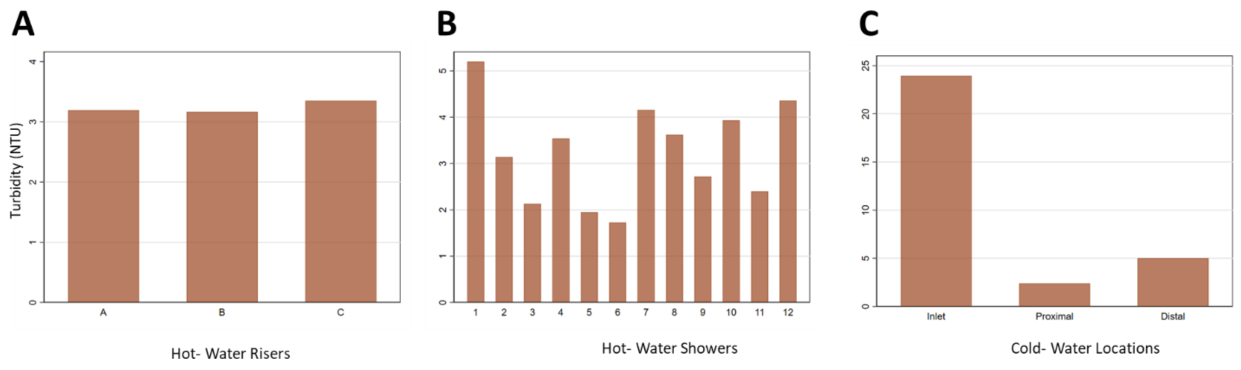


**Figure S6.** A) Mean turbidity across risers from hot-water samples. B) Mean turbidity across 12 showers from hot-water samples. C) Mean turbidity across cold-water sample locations.

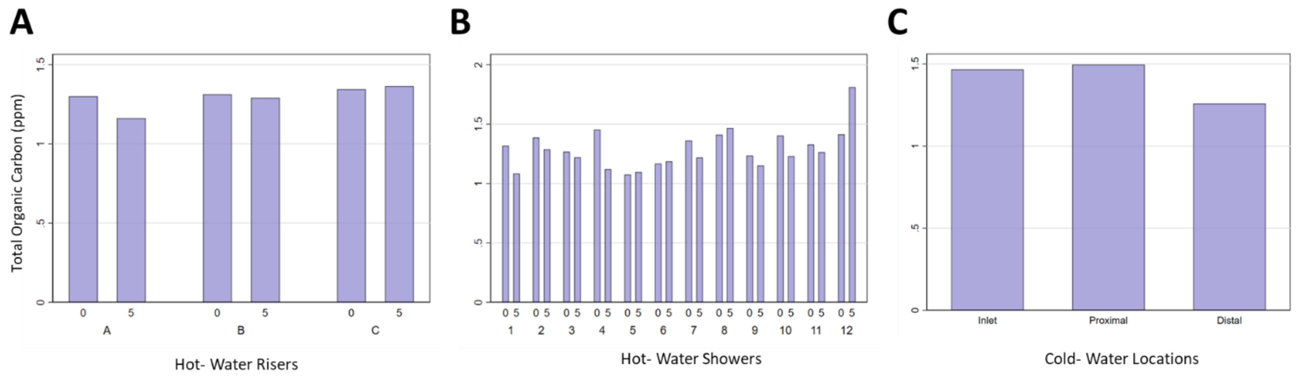


**Figure S7.** A) Mean TOC across risers from hot-water samples. B) Mean TOC across 12 showers from hot-water samples. C) Mean TOC across cold-water sample locations. “0” indicates Time 0 and “5” indicates Time 5.
